# Deep learning for subtypes identification of pure seminoma of the testis

**DOI:** 10.1101/2022.05.16.22275153

**Authors:** Kirill E. Medvedev, Paul H. Acosta, Liwei Jia, Nick V. Grishin

## Abstract

The most critical step in the clinical diagnosis workflow is the pathological evaluation of each tumor sample. Deep learning is a powerful approach that is widely used to enhance diagnostic accuracy and streamline the diagnosis process. In our previous study using omics data, we identified two distinct subtypes of pure seminoma. Seminoma is the most common histological type of testicular germ cell tumors (TGCTs). Here we developed a deep learning decision making tool for the identification of seminoma subtypes using histopathological slides. We used all available slides for pure seminoma samples from The Cancer Genome Atlas (TCGA). The developed model showed an area under the ROC curve of 0.896. Our model not only confirms the presence of two distinct subtypes within pure seminoma but also unveils the presence of morphological differences between them that are imperceptible to the human eye.

## Introduction

Testicular seminoma is the most prevalent histological subtype of testicular germ cell tumors (TGCT), accounting for the highest incidence rate among all types of testicular cancer ^1^. TGCTs are the most frequent type of solid cancer affecting men between the ages of 15 and 44 ^1^ and rank second among adult cancers in terms of life years lost per person dying of cancer ^2^. The treatment protocol for seminoma typically includes orchiectomy followed by either platinum-based chemotherapy utilizing cisplatin or radiation therapy ^3^. While current treatments for seminoma have high efficacy and survival rates for patients, they also carry the risk of around 40 severe and potentially life-threatening long-term side effects, such as infertility, neurotoxicity, hypercholesterolaemia, secondary cancers and death ^4^. The presence of elevated platinum concentrations from chemotherapy in the bloodstream can persist at levels up to 1,000 times higher than the norm for a duration of 20 years, potentially contributing to various long-term effects ^5^. Prolonged exposure to elevated platinum levels can result in vascular damage and is highly likely to be linked with the onset of neuropathy ^6^ and cardiovascular diseases ^7^. After undergoing chemotherapy, patients with TGCT exhibited a decrease of 3.6 dB in hearing for each additional 100 mg/m2 of cumulative cisplatin dose ^8,9^. Relapse occurs in approximately 20% of seminoma cases, and the underlying reasons for this phenomenon remain unclear ^10^, however there are several well-known seminoma risk factors such as rete testis, lymphovascular invasion, cryptorchidism, mutations in KRAS and KIT genes ^11,12^. Patients experiencing a relapse will receive further treatment involving chemotherapy and radiation therapy, which intensify the side effects considerably. Recently we discovered two distinct subtypes of pure seminoma of the testis based on omics data ^13,14^. Two identified seminoma subtypes revealed significant differences in the rates of loss of heterozygosity, the level of expression of lncRNA associated with cisplatin resistance, the activity of double stranded DNA breaks repair mechanisms and the pluripotency stage. Seminoma subtype 1 exhibits a higher pluripotency state, while subtype 2 reveals attributes of reprogramming into non-seminomatous lineages of TGCT, which are more aggressive and require higher dose of chemotherapy drugs ^3^. We showed that subtype 1 of seminoma, which is less differentiated, exhibits an immune microenvironment characterized by a significantly lower immune score and a larger fraction of neutrophils ^14^. These features are indicative of the immune microenvironment at an early developmental stage. Moreover, subtype 2 revealed the overexpression of genes related to the senescence-associated secretory phenotype, which might be one of the reasons for seminoma immunotherapy failure ^14^. Therefore, we suggested that seminoma subtype 2 might require an adjustment to its treatment strategy. The development of subtype-specific therapy for seminoma can reduce the risk of chemotherapy overtreatment in TGCT patients and enhance the quality of life for TGCT survivors.

Deep learning (DL) is a powerful tool capable of extracting previously hidden information directly from routine histopathology images of cancer tissue, simplifying, speeding up, and automating clinical decision-making ^15^. The performance of modern DL methods applied to pathological data often exceeds that of human pathologists ^15^. Most pathologists work under conditions of an extreme work overload ^16^. An overworked pathologist can result in the misinterpretation of pathological data that affects patients’ health and quality of life. DL applications aim to simplify and speed up routine pathological workflows and reduce pathologists’ overload burden.

Here, we have developed a DL-based approach to examine potential histopathologic differences between seminoma subtypes that were previously identified using omics data. Additionally, our goal is to utilize this approach to detect and classify these subtypes based on histopathological slides. Our findings demonstrate that pure seminoma subtypes cannot be classified solely based on histopathological features. However, the developed DL-based model revealed histopathological differences between these subtypes, as indicated by the area under the ROC curve (AUC) values.

## Materials and Methods

### Data set preparation

We used all hematoxylin and eosin (H&E) histopathological slides available at The Cancer Genome Atlas (TCGA) data portal (TCGA-TGCT study) for 64 pure seminoma, which comprised 156 whole slide images (WSIs). Based on our previous study, we assigned 40 out of 64 samples to seminoma subtype 1 (101 WSIs) and the remaining 24 samples to seminoma subtype 2 (55 WSIs) ^13^. Pure seminoma regions of interest (ROIs) were designatedand verified for each WSI by a genitourinary specialized pathologist using Aperio ImageScope version 12.1. During our analysis of pure seminoma H&E slides from the TCGA portal, we identified two samples (TCGA-2G- AAG9, TCGA-2G-AAH0), initially reported as pure seminoma. However, upon further examination, they should be reclassified as mixed GCT since they contain other types of GCT (teratoma and embryonal carcinoma) in addition to seminoma (Fig.1). Consequently, these cases were removed from our data set.

**Figure 1.**
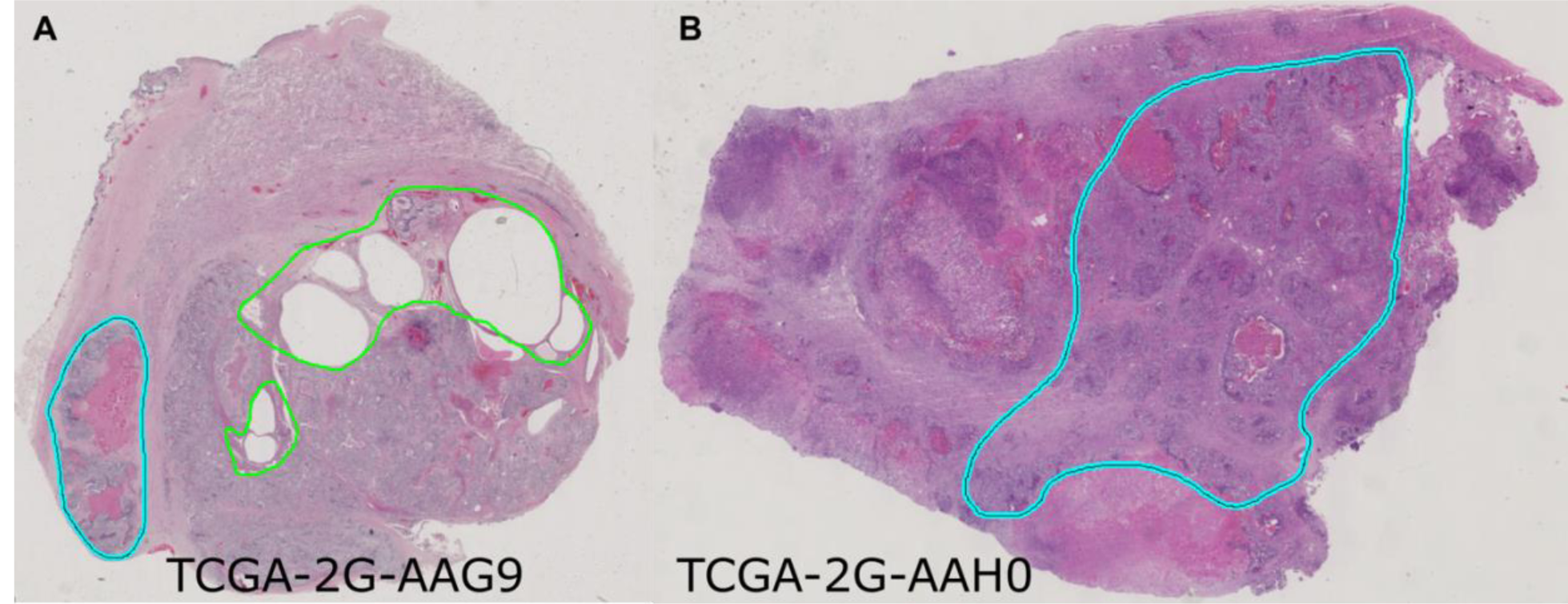
Slide images of samples containing additional types of GCT. (A) **TCGA-2G-AAG9. (B)** TCGA-2G-AAH0. Teratoma tissue is shown in green, embryonal carcinoma in cyan.

Verified ROIs were subsequently split into smaller tiles (300x300 pixels) at a 20X magnification with a 50% overlap using DeepPath package ^17^ (Fig. 2). Tiles that contained more than 20% of background were removed. We conducted a manual check and excluded tiles of poor quality that contained out of focus images and defects, such as scratches, dirt and folded tissue. TGCTs are relatively uncommon compared to other cancers ^2^, and therefore histopathological data availability is limited. Thus, image data augmentation technique was applied to the dataset of tiles to create synthetic variations of the images and expand the training dataset. We used the following augmentations from the Imgaug library ^18^: random rotation by 0°, 90°, 180°, 270°, random increase and decrease of contrast, brightness and saturation.

**Figure 2.**
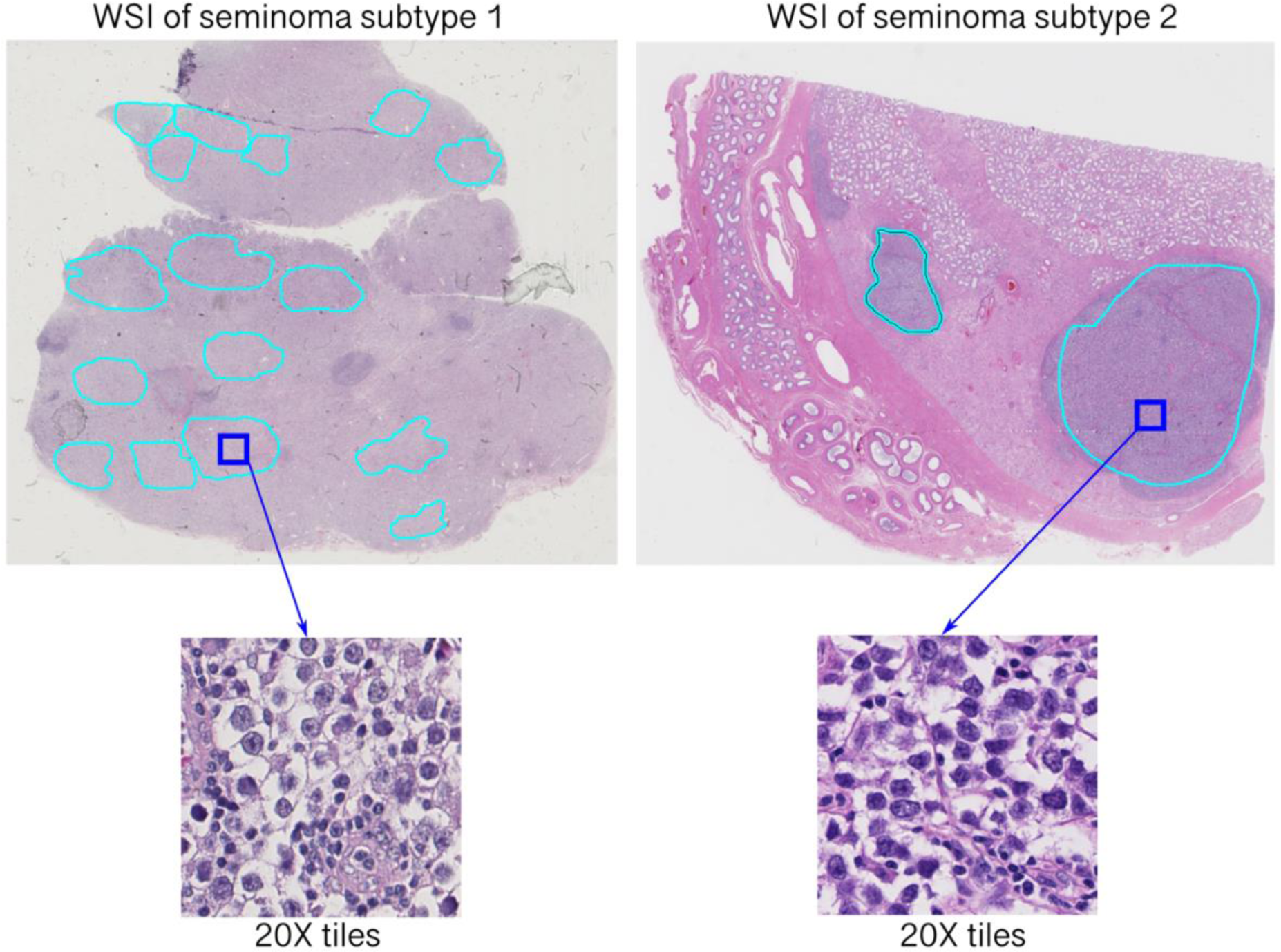
Extraction of tiles from annotated whole slide images (WSI) of two subtypes of pure seminoma.

### Training the model

We conducted all computational experiments at the BioHPC computing facility (Lyda Hill Department of Bioinformatics, UT Southwestern Medical Center, TX, USA). TensorFlow package ^19^ version 2.5.0 was used for developing DL model based on Convolution Neural Networks (CNNs) method. The software stack for GPU acceleration included CUDA 11.2 and cuDNN 8.1. We employed a convolutional neural network with MobileNet ^20^ architecture that includes 85 convolutional layers. During the training top 29 convolutional layers were kept frozen (fixed). We used sigmoid activation and the Binary Cross Entropy loss function, which is used when there are only two label classes (seminoma subtypes 1 and 2). The Adam optimizer was selected due to its superior performance in terms of both speed of convergence and accuracy ^21^ with a learning rate of 0.001. Due to imbalanced dataset we used subtype (class) weights calculated as follows: w_1_ = (1 / s_1_) * (total / 2.0) and w_2_ = (1 / s_2_) * (total / 2.0), where s_1_, s_2_ – number of tiles for subtypes 1 and 2, and total is overall number of tiles. The neural network was initialized from ImageNet-pretrained ^22^ weights. Model training was performed for 20 epochs with 3-fold cross-validation. Tiles belonging to a particular sample were included only in one subset of data – either training or validation.

### Statistical analysis

We used the area under receiver operating characteristics (ROC, AUC) curve and accuracy as evaluation metrics to measure the tile-level and sample-level performance of the developed model. The ROC curve was defined as false-positive rate (1-specificity) on the x-axis versus true positive rate (sensitivity or recall) on the y-axis. Specificity = TN / (TN+FP), Sensitivity = TP / (TP+FN), Accuracy = (TP+TN) / (TP+FP+TN+FN), where FP, FN, TP and TN are false positives, false negatives, true positives and true negatives, respectively.

### Nuclei segmentation

Nuclei segmentation of seminoma tiles was conducted using TIA Toolbox 1.4.1 ^23^. We applied the HoVer-Net model ^24^ that has been already trained on the PanNuke dataset ^25^ and incorporated in the TIA Toolbox. The calculation of nuclei size was performed using the Python library scikit- image ^26^.

## Results and Discussion

Overview of the whole experiment is shown on Figure 3. The performance of the model was evaluated using the area under the ROC curve metric (Fig. 4B) and confusion matrices (Fig. 4C). The developed model showed highest AUC = 0.896 (Fig. 4B). Trained model for identification of pure seminoma subtypes is available in open access at GitHub (https://github.com/kirmedvedev/seminoma-subtypes).

**Figure 3.**
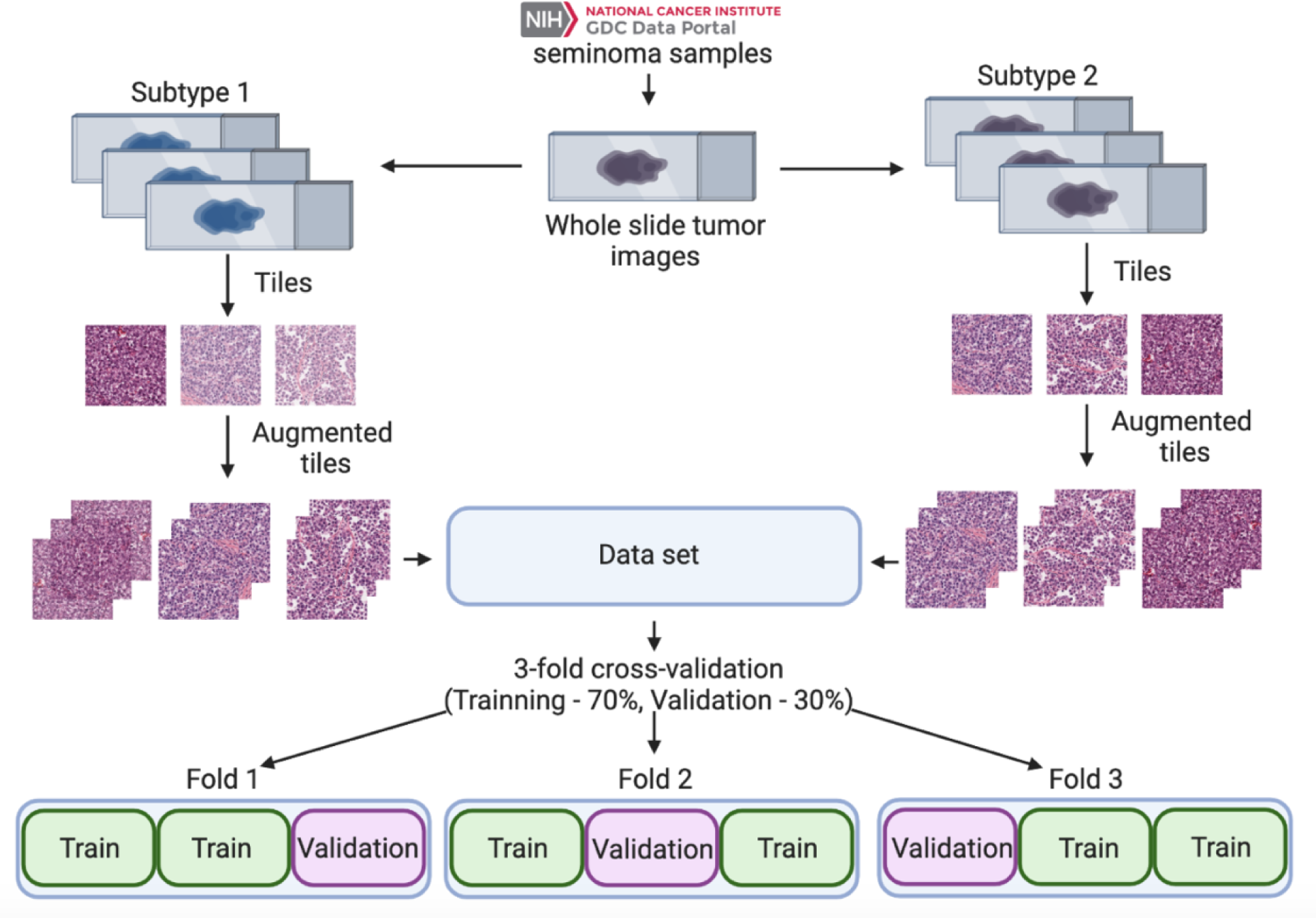
**Overview of dataset preparation, training and validation process.**

**Figure 4.**
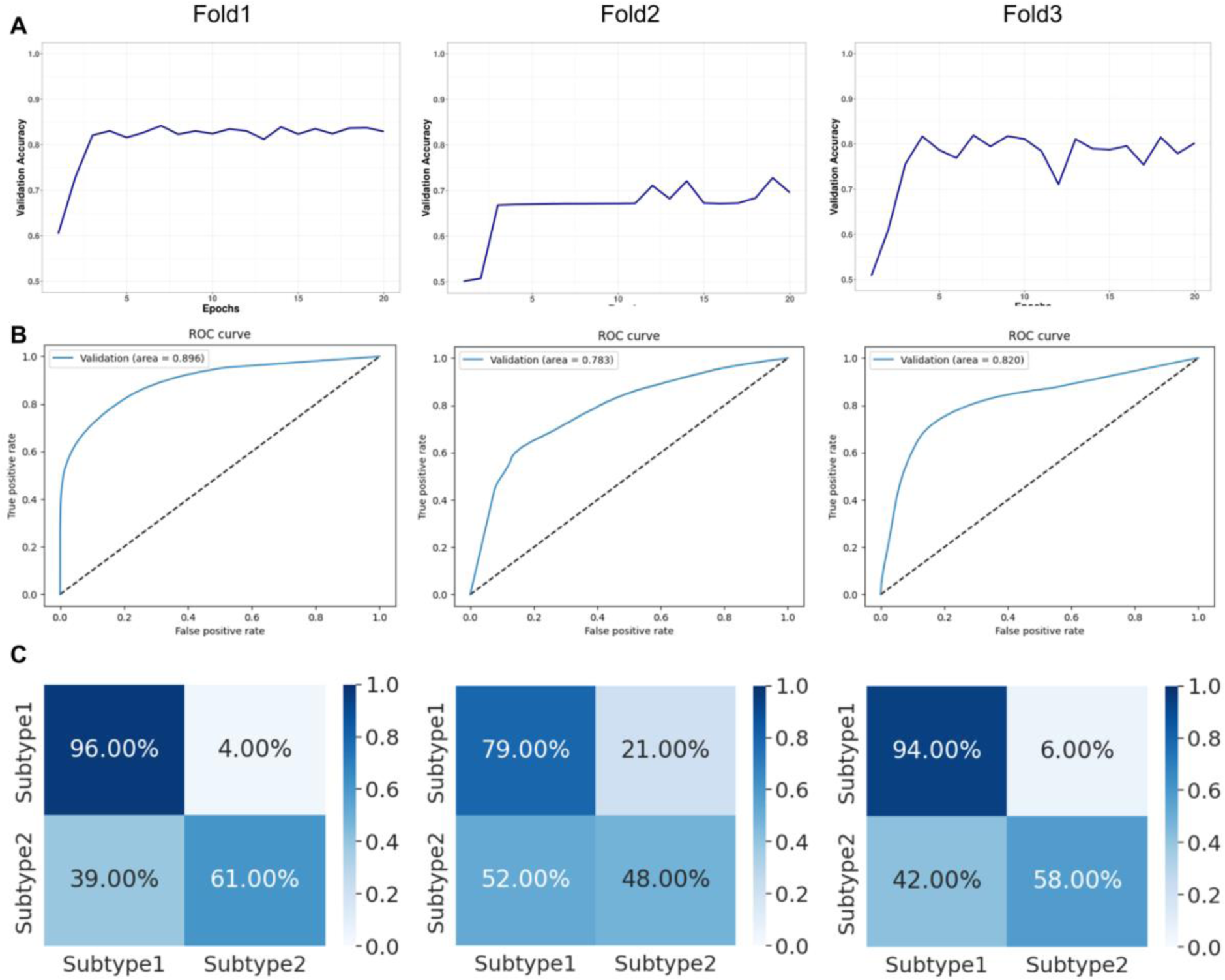
Validation statistics. (A) Validation accuracy. **(B)** Receiver operating characteristic (ROC) curves for validation set. **(C)** Normalized confusion matrices.

Every sample of solid tumor undergoes a detailed analysis by a professional pathologist, which includes verifying the presence of cancer tissue and annotating tumor regions. This is a crucial process in the clinical diagnosis routine. Inaccuracies in pathology reports can critically affect the quality of patient care. An audit of pathology reports showed that if a sample size of 50 gives a sample error rate of 2% there is a 95% probability that the true error rate is up to 10.9% ^27^. Moreover, up to one-third of clinicians do not always understand pathology reports, leading tomisinterpretation and uncertainty in clinical diagnosis ^28^. DL approaches applied to histopathological slide images aim to speed up the diagnosis significantly and simplify their implementation into the clinical workflow. TGCTs and seminoma histopathology images, in particular, have not been extensively studied using DL method, and very limited studies are available nowadays. DL approached were previously applied to TGCTs WSIs for detecting tumor-infiltrating lymphocytes ^29^, detecting lymphovascular invasion ^30^ and developing tumor/normal classifier ^31^.

In this report, we present our first version of the DL decision making tool for the identification of pure seminoma subtypes using histopathological slides. We hypothesize that considering seminoma subtypes during the development of a treatment strategy may improve its clinical management, and the implementation of the developed model will enhance diagnostic accuracy and reduce potential errors. This is especially crucial when subtypes cannot be distinguished by a pathologist which is the case with pure seminoma. The developed model showed the capability to distinguish pure seminoma subtypes (Table 1),confirming our previous findings ^13,14^. This also indicates the presence of morphological differences between seminoma subtypes. We believe that the morphological differences may be due to the difference in the immune microenvironment between the two subtypes. Previously, using deconvolution methods for bulk RNA-seq data of the seminoma subtypes from TCGA, we showed that the neutrophil fraction is significantly higher for subtype 1 ^14^. Moreover, according to TCGA clinical data, seminoma subtype 2 revealed an increased occurrence of lymphovascular invasion, with a rate of 43% compared to 25% for subtype 1. We also conducted nuclei segmentation and calculated nuclei sizes for both subtypes. Our analysis revealed no significant differences in nuclei size distributions between the seminoma subtypes (Fig. 5A, B).

**Figure 5.**
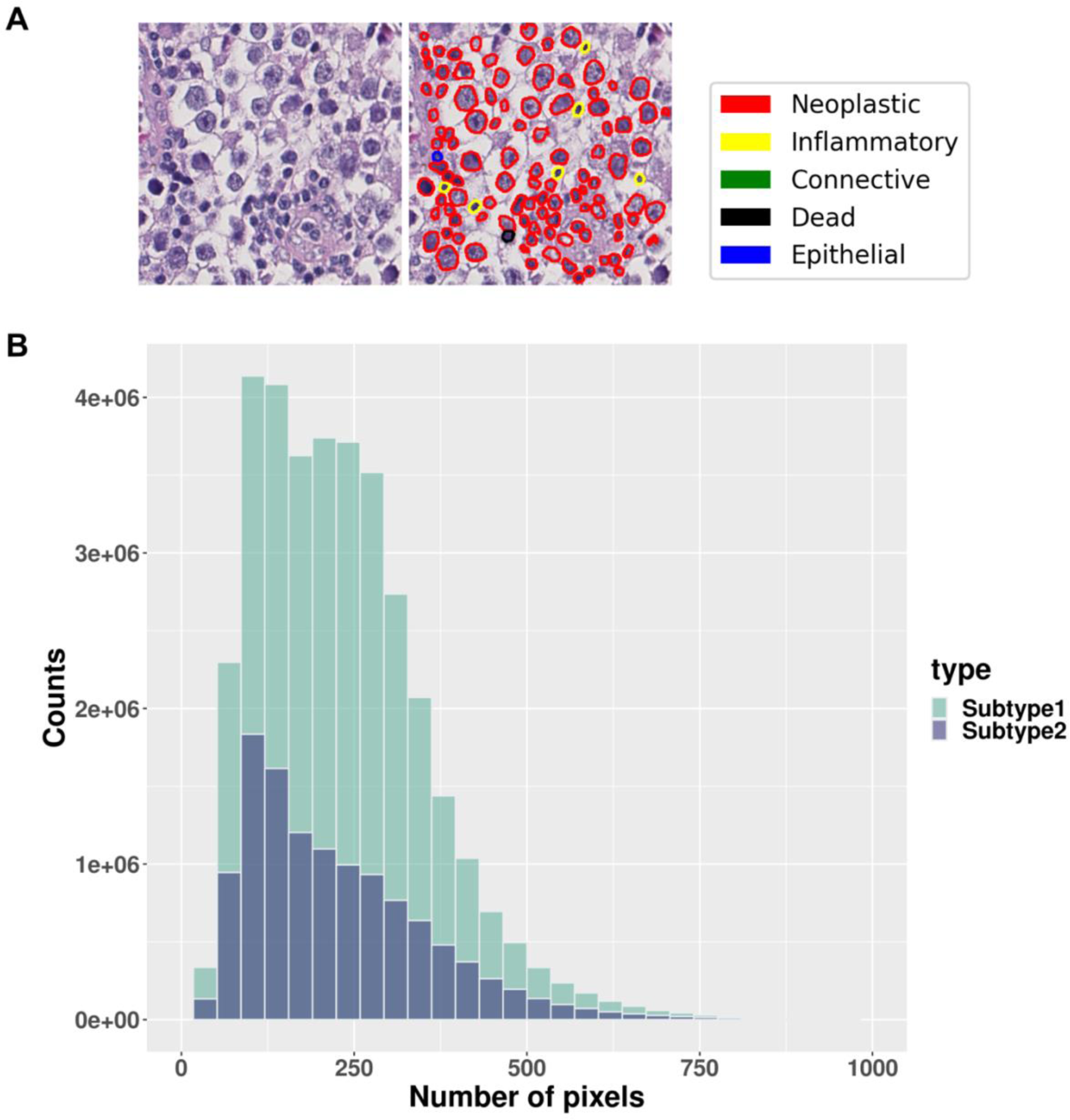
Nuclei segmentation results. (A) Visualization of nuclei segmentation results. **(B)** Comparison of seminoma cell nuclei size distribution between two subtypes.

**Table 1.** Prediction of seminoma subtypes using developed DL model. S1 – subtype 1, S2 – subtype 2.

| Sample ID | Transcriptomic subtype | DL model prediction | Sample ID | Transcriptomic subtype | DL model prediction |
| --- | --- | --- | --- | --- | --- |
| TCGA-XY-A9T9 | S1 | S1 | TCGA-2G-AAH3 | S1 | S1 |
| TCGA-WZ-A7V4 | S1 | S1 | TCGA-SB-A6J6 | S1 | S1 |
| TCGA-2G-AAEX | S1 | S1 | TCGA-2G-AAHP | S1 | S1 |
| TCGA-2G-AAF6 | S1 | S1 | TCGA-VF-A8AB | S2 | S1 |
| TCGA-S6-A8JX | S1 | S1 | TCGA-ZM-AA05 | S2 | S2 |
| TCGA-XY-A89B | S1 | S1 | TCGA-XE-AANR | S2 | S2 |
| TCGA-2G-AAFG | S1 | S2 | TCGA-ZM-AA0D | S2 | S2 |
| TCGA-2G-AAH8 | S1 | S1 | TCGA-2G-AAHN | S2 | S2 |
| TCGA-2G-AAF1 | S1 | S1 | TCGA-4K-AAAL | S2 | S1 |
| TCGA-WZ-A7V3 | S1 | S1 | TCGA-VF-A8AE | S2 | S1 |
| TCGA-S6-A8JY | S1 | S1 | TCGA-ZM-AA0B | S2 | S2 |
| TCGA-VF-A8AA | S1 | S1 | TCGA-XE-AAOF | S2 | S1 |
| TCGA-WZ-A7V5 | S1 | S1 | TCGA-ZM-AA0F | S2 | S2 |
| TCGA-SO-A8JP | S1 | S1 | TCGA-2G-AAHA | S2 | S1 |
| TCGA-XE-A8H4 | S1 | S1 | TCGA-2G-AAHT | S2 | S1 |
| TCGA-XE-A8H5 | S1 | S1 | TCGA-4K-AA1H | S2 | S2 |
| TCGA-XE-A9SE | S1 | S2 | TCGA-VF-A8A9 | S2 | S2 |
| TCGA-XE-AANJ | S1 | S1 | TCGA-VF-A8AC | S2 | S1 |
| TCGA-XE-AANV | S1 | S1 | TCGA-XE-AAO6 | S2 | S2 |
| TCGA-XE-AAO3 | S1 | S1 | TCGA-XE-AAOL | S2 | S2 |
| TCGA-YU-A90Q | S1 | S2 | TCGA-YU-A90S | S2 | S1 |
| TCGA-YU-A90W | S1 | S1 | TCGA-ZM-AA06 | S2 | S2 |
| TCGA-YU-A912 | S1 | S2 | TCGA-ZM-AA0E | S2 | S2 |
| TCGA-S6-A8JW | S1 | S1 | TCGA-ZM-AA0H | S2 | S1 |
| TCGA-2G-AAEW | S1 | S1 | TCGA-ZM-AA0N | S2 | S2 |
| TCGA-2G-AAF4 | S1 | S1 | TCGA-2G-AAHL | S2 | S1 |
| TCGA-4K-AA1I | S1 | S2 | TCGA-4K-AA1G | S2 | S2 |
| TCGA-2G-AAFE | S1 | S1 |  |  |  |

However, the accuracy of identifying subtype 1 samples by the developed model is higher than of subtype 2 samples. This could be due to certain limitations of the model. First, the current model was developed using a limited dataset. Second, only one architecture type of CNN was tested. In future work, this model should be verified using an expanded dataset and several addition CNN architecture types.

## Conclusion

In this study we developed a DL-based model to investigate the presence of histopathological distinctions between two previously identified subtypes of pure seminoma, which were initially characterized using omics data. The objective was to provide further evidence supporting the existence of seminoma subtypes. The results of our analysis revealed histopathological differences between the two subtypes of pure seminoma. These findings provide additional confirmation and support the notion that seminoma can be further stratified into distinct subtypes. These results highlight the potential of histopathological analysis as a complementary tool in subtype classification, offering additional insights alongside other omics-based approaches. However, our study also provides evidence suggesting that pure seminoma subtypes cannot be reliably classified based solely on histopathological features. Despite the observed histopathological differences between the subtypes, these distinctions alone are not sufficient for accurate subtype classification.

## Data Availability

All data produced are available online at https://github.com/kirmedvedev/seminoma-subtypes

## Acknowledgements

Authors are grateful to Dr. Satwik Rajaram for helpful discussions. Authors are grateful to TCGA data portal for providing access to TGCT datasets. This research was supported in part by the computational resources provided by the BioHPC computing facility located in the Lyda Hill Department of Bioinformatics, UT Southwestern Medical Center, TX. URL: https://portal.biohpc.swmed.edu

## Ethics statement

Not applicable.

